# Socioeconomic Disparities and COVID-19 Vaccination Acceptance: Experience from Israel

**DOI:** 10.1101/2021.01.28.21250716

**Authors:** Gil Caspi, Avshalom Dayan, Yael Eshal, Sigal Liverant-Taub, Gilad Twig, Uri Shalit, Yair Lewis, Avi Shina, Oren Caspi

**Affiliations:** The Faculty of Electrical Engineering the Technion, Israel; the B. Rappaport Faculty of Medicine, Technion, Haifa, Israel; Division of General Medicine, Medical Directorate, Israel Ministry of Health, Jerusalem, Israel; The Israeli Defense Forces Medical Corps; The Dr Pinchas Bornstein Talpiot Medical Leadership program Sheba Medical Center, Ramat Gan Israel; Sackler School of Medicine, Tel-Aviv University, Israel; Institute of Endocrinology Diabetes and Metabolism, Sheba Medical Center, Tel Hashomer, Israel; Department of Military Medicine, The Hebrew University, Jerusalem, Israel; The Faculty of Industrial Engineering and Management, Israel; Maccabi Healthcare Services, Israel; The department of Cardiology, Rambam Health Care Campus

## Abstract

COVID-19 vaccination acceptance has a key role in mitigating the pandemic. Concern has been raised that vaccination rates will be limited in demographically defined areas of lower income. Israel’s rapid vaccination campaign may allow to assess these assumptions in real-world and to devise tools for effectively focusing the vaccination efforts. We analyzed the correlation between COVID-19 vaccination rates, socioeconomic status (SES) and active COVID-19 disease burden. We carried out a nationwide study, based on data provided by Ministry of Health of COVID-19 vaccination rates in all municipalities in Israel up to January 12^th^, 2021. Municipal Vaccination rates of population older than 60 significantly correlated with the socioeconomic status (r=0.83, 95% confidence interval [0.79 to 0.87]). Finally, we established a novel metric for focusing the vaccination efforts based on % vaccinations and active disease burden. In Israel, a case-model country for COVD-19 vaccinations, vaccination rates were strongly correlated with SES. The study findings demonstrate the need to directly target vaccination acceptance to socio-economically disadvantaged populations and suggest potential tools for policymakers to focus their efforts.

## Introduction

The coronavirus disease 2019 (COVID-19) pandemic has resulted in the mortality of over 2 million people worldwide and has become a leading cause of death in adults^1^. The rapid development of COVID-19 vaccines provides new hopes regarding our ability to battle the pandemic^2-4^. However, while the approval and availability of COVID-19 vaccinations is progressing around the globe, the pandemic is reaching a calamitous scale and overwhelming health care systems capabilities. Therefore, the limited roll-out of vaccinations relative to the COVID-19 burden mandates prioritization of the vaccination effort^5^. Moreover, prior survey studies suggested that vaccination rates may be diminished in among demographically defined groups with lower education and income levels^6^. Currently, Israel, a highly heterogenous country, leads the global effort for vaccinating its population making it a suitable case study for other countries as they form their vaccination strategies^7^. Many countries adopted strategies based on prioritizing elderly people as they are at the greatest risk for severe COVID-19 related disease and mortality. In Israel, people over the age of 60, together with healthcare personnel entrenched in the disease frontlines, were thus the first to be offered the vaccine^8^.

Elderly people, at greatest risk for COVID-19 related mortality and morbidity, from municipalities with a lower socioeconomic status (SES) may suffer from lower availability of healthcare resources, leading to poorer health and lower acceptance of public healthcare measures such as vaccines^9^. Concurrently, some of these populations at low SES and of rural locations are more severely affected by the COVID-19 pandemic in Israel and worldwide^10-12^. The overarching hypothesis of our research is that population with a lower SES are subject to a double hit risk in the setting of the current pandemic, a higher rate of contagious disease coupled with a lower rate of vaccination

## Methods

### Study population

<u>All residents in Israel enjoy medical coverage by law, provided by four health maintenance organizations (HMOs). All HMOs use electronic medical records and provide COVID-19 related data and vaccination information to the Israeli Ministry of Health. The study population consisted of nationally anonymized and aggregated data provided by the Ministry of Health.</u>

### Data Preparation

#### The Socioeconomic status

The SES scoring was based on the Israeli Central Bureau of Statistics (ICBS) scoring system. Accordingly, each place of residence, obtained from the Israeli Ministry of Interior, is ranked from lowest to highest SES. The score stratifies all municipalities into 10 decile groups, according to multiple variables that might affect SES, such as age distribution, level of unemployment, and available work force, education (the proportion of undergraduate students and those entitled to a high school diploma), average income per capita, and the proportion that receives income support. The SES data were processed from ICBS SES report 2017 [https://www.cbs.gov.il/he/mediarelease/doclib/2020/403/24_20_403t1.xlsx] and the rank of each municipality was used. The population data by age group in each municipality 2019 [https://www.cbs.gov.il/he/publications/doclib/2017/population_madaf/population_madaf_2019_1.xlsx] was taken from ICBS. Given that municipality’s population was updated two years ago, in some extreme cases the number of vaccinated persons over the age of 60 (currently counted) is higher than that found in the municipality’s population.

#### COVID-19 Vaccination rates and active disease burden

Data of active COVID-19 cases for each municipality was derived from the municipalities table in COVID-19 database of the Israeli Ministry of Health (MOH) [https://data.gov.il/dataset/covid-19]. First dose vaccinations by municipality and age group data were derived from a status report published by the Israeli MOH on the 13^th^ of January 2021 [https://data.gov.il/dataset/covid-19/resource/12c9045c-1bf4-478a-a9e1-1e876cc2e182]. Only people aged>16 were deemed eligible to be vaccinated. In the provided data by the Israeli MOH, values between 1 to 14 cases/vaccinations in a municipality were marked as ‘<15’ without indicating the exact number, due to privacy requirements. In those cases, missing data imputation was performed by replacing ‘<15’ with the lower bound of 1.

The active cases and vaccination data were reported in 279 municipalities in Israel. SES was reported in 196 out of the 279 municipalities. Vaccinations of people over the age of 60 were aggregated out of 4 age groups (‘60-69’, ‘70-79’, ‘80-89’, ‘90+’). We filtered out the municipalities that had more than 1 sub-group with missing data (‘<15’) – and accordingly a total of 12 municipalities were filtered out. Hence, our analysis was performed using data of 183 municipalities in Israel. Color coding of municipalities was conducted by ranking the relevant metric according to percentiles. The lower 20 and upper 80 percentile were color-coded in red and green, respectively. Color coding was spectrally determined according to the relevant percentile.

#### The VNR

To characterize the association between the COVID-19 active case burden, and the vaccination rates of the population older than 60, we devised the vaccination need ratio (VNR). VNR is calculated by dividing the total number of active cases (per 10,000 people) by the rate of vaccination of the population over 60 in a given municipality (m)

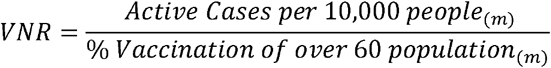

We then examined the association between the municipal SES to the VNR. Finally, we color-coded the municipalities according to the VNR metric where municipalities at top 20 percentiles of VNR were coded in red (*e*.*g*. high number of active cases per 10,000 people and low rates of vaccination) and those in the bottom 20 percentiles were coded in green with range spectral coding of all other municipalities, accordingly. We applied our findings to create a heatmap.

#### Statistical analysis

The correlation was analyzed by using a weighted Pearson correlation (according to the municipality population over 60 from the total population over 60 from the evaluated municipalities) and the confidence interval was calculated using a bootstrapping method with an alpha of 0.95. Basic arithmetic calculations were conducted using Python Pandas and NumPy. Statistical analysis was done using Python SciPy.

#### Data Availability

Map data are copyrighted by Mapbox contributors and are available from https://vaccinations.covid19maps.org/

## Results

To assess whether the vaccination rates were related to SES, we evaluated the correlation between SES rank and vaccination rates in the at-risk population (aged over 60). Vaccination rates strongly correlated with SES municipality (r=0.83, 95% confidence interval [0.79 to 0.87]) as shown on Figure 1. Assessment of the correlation between the burden of total active cases (per 10,000 people) in a municipality and the vaccination rate of population over 60, identified a significant negative correlation (r=-0.47, 95% confidence interval [-0.59 to −0.30]) as displayed in Figure 2.

**Figure 1:**
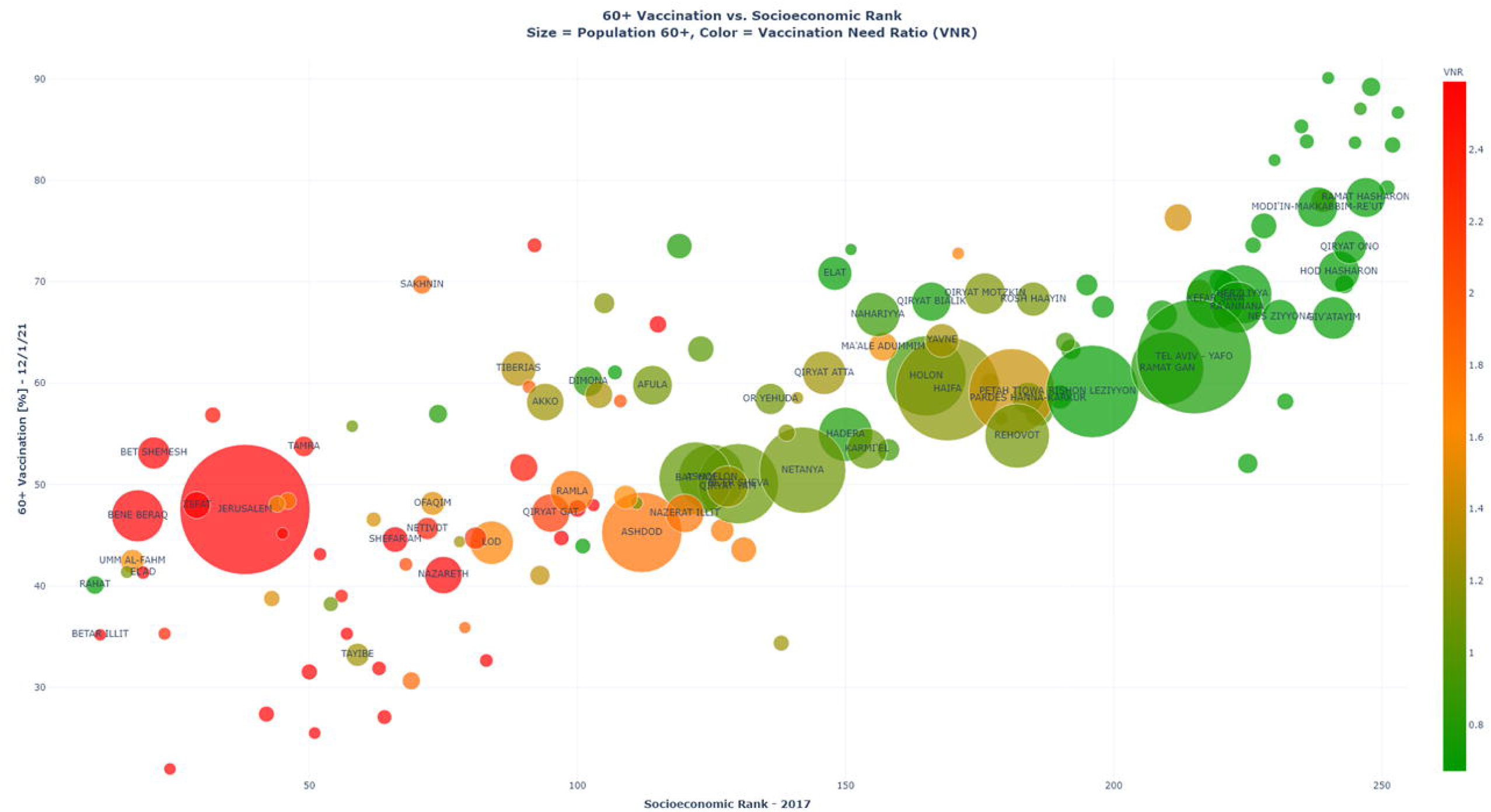
Correlation between vaccination of at-risk population and Socioeconomic status. A bubble chart depicting the association between the percentage of the vaccinated population over the age of 60 in a municipality and the municipality socioeconomic status (SES) as ranked by the Israeli Central Bureau of Statistics. A strong positive correlation (r=0.83, 95% confidence interval [0.79 to 0.87]) was found. The Bubble size is indicative of the size of the population than 60 while the spectral coding represents the vaccination need ratio-VNR (columns on the righthand side). Municipalities with a population older than 60 of less than 1,000 are not displayed and only those with a total population of more than 30,000 are named in the chart.

**Figure 2:**
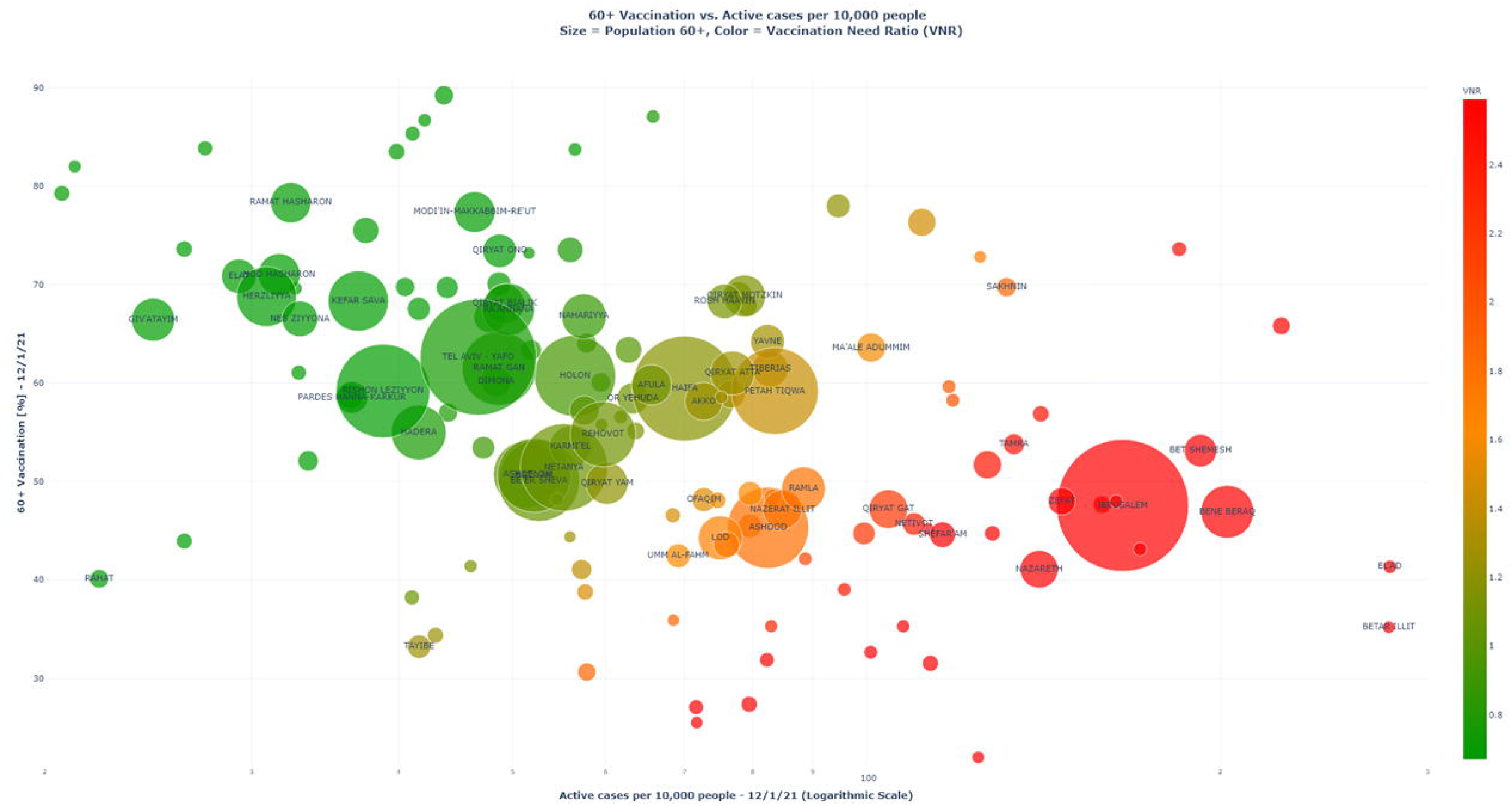
Correlation between vaccination of at-risk population and active COVID-19 cases. A bubble chart depicting the association between the percentage of vaccinated population over the age of 60 and the active COVID-19 cases (on a logarithmic scale) to the rate of the in each municipality. A moderate negative correlation (r=-0.47, 95% confidence interval [-0.59 to −0.30]) was found. The Bubble size is indicative of the size of the population older than 60 while the spectral coding represents the vaccination need ratio-VNR (column on the righthand side). Municipalities with a population older than 60 of less than 1,000 people are not displayed and only those with a total population of more than 30,000 people are named in the chart.

The record levels of COVID-19 cases mandates harnessing the vaccination efforts as a mitigation tool. To further focus the vaccination efforts, beyond the prioritization based on age, we established the vaccination need ratio (VNR). The VNR aid in highlighting municipalities with high active COVID-19 case burden and low vaccination rates. Importantly, we identified a significant negative correlation between VNR and SES (r=-0.80, 95% confidence interval [-0.88 to −0.66]), further supporting the need to directly target the vaccination efforts to socio-economically disadvantaged populations.

To assess the geographical dispersion of the VNR across the country, we generated a color-coded heatmap that portrays the VNR ranges between different municipalities (Figure 3). The generated map demonstrated high VNR in northern Israel, the seam zones and municipalities heavily populated with minorities.

**Figure 3:**
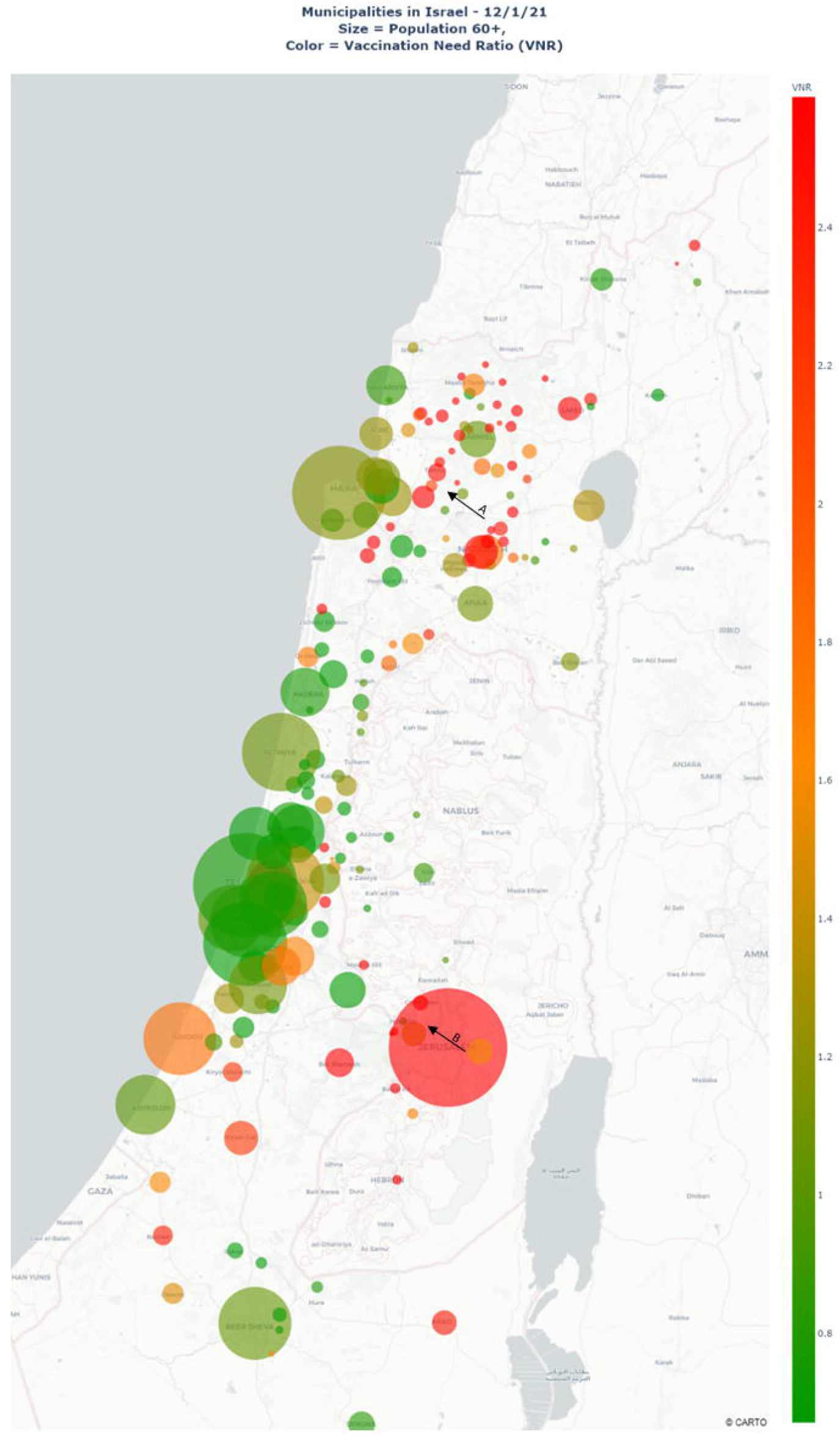
Municipal nation-wide vaccination need ratio-VNR heat map. A heat map of Israel portraying the municipal nation-wide vaccination need ratio-VNR (color-coded) of the different municipalities. Arrow A and arrow B point to municipalities in Northern Israel and the seam zone, respectively. Those municipalities have a VNR, which might be indicative of a forming geographical pocket of low immunity. The Bubble size is indicative of the size of the population over the age of 60.

## Discussion

The detrimental prices inflicted by the COVID-19 pandemic necessitate rapid and effective vaccinations of all at-risk populations across the spectrum of all socioeconomic backgrounds to prevent severe morbidity and mortality while reducing the disease burden on health care systems. Israel employs a rapid and robust vaccination effort and as of 29^th^ of January has the highest percentage of vaccinated population, making it a case study for other nations worldwide. In the current analysis, we identified a significant correlation between municipal vaccination rates and socioeconomic status. Also, we demonstrated that municipalities with a high current COVID-19 disease burden had a lower rate of vaccination of the elderly population. The metric established, the vaccination need ratio (VNR), was found to strongly correlate with the municipal SES. Finally, we generated a heatmap highlighting areas of high VNR, allowing us to outline the need for vaccination in a geographical context and assisting policymakers in avoiding pockets of high COVID-19 morbidity and low immunity.

We found that municipalities with a lower SES, often populated with ethnic minorities in Israel, suffer from a higher disease burden, yet their at-risk population has not been vaccinated against COVID-19 in the targeted rates. In Israel, like other countries, populations from lower SES and minorities suffer from lower accessibility and availability to healthcare resources as well as cultural barriers. This leads to a reduced willingness to actively partake in recommended public health measures (social distancing, compliance to mask-wearing), putting these populations at an increased risk for COVID-19 infection during the pandemic. These socioeconomic gaps may be overemphasized given the introduction of a vaccine based on novel mRNA technology that may raise objection and safety concerns.

The main strength of our study is due to the nationwide availability of COVID-19 aggregated data of an ethnically heterogenous population thus making our findings generalizable to other countries. Second, the timeliness of our findings is of temporal relevance and our analysis was conducted prior to initiation of the protective effects of the vaccines expected to further widen the differences between municipalities with low and high SES. Third, a national diagnostic effort (a rate of over 12 tests/day per 1,000 people) coupled with real-time monitoring of the vaccination rates allow for a timely and accurate derivation of the VNR scoring system. The national data is dynamically updated on our website (https://vaccinations.covid19maps.org/) enabling policymakers to track the effect of their actions.

Our findings have limitations. The SES is calculated differently in each country, though the national ICBS classification correlative with metabolic disease^13^ making our findings generalizable. Also, in this study it is used as an ecological variable and is not indicative of individual health, yet this is a more appropriate representation of its implication on municipal vaccination. We did not have available data regarding active disease burden of population over 60, or the disease severity of active cases within municipalities. As data were aggregated we did not have available personal data regarding COVID-19 risk factors, though older age is the most important risk factor for severe COVID-19^14^. Finally, we were not able to remove the population aged over 60 who recovered or died from COVID-19 from the vaccination potential population in each municipality, though Israel has suffered from approximately 4,000 COVID-19 related deaths only, with approximately 5% of the population recovered from the disease hence this does not affect our findings. These data are an interim analysis based on the rates of the first dose of vaccination, and data of 2^nd^ vaccination shot compliance are not yet available. The analysis time point, conducted 3 weeks following the initiation of vaccinations in Israel, aims at evaluating vaccination rollout and distribution as well as compliance and does not intend to evaluate vaccination efficacy in mitigating the pandemic. Moreover, as the mRNA vaccine logistics mandate unique temperature control and careful mobilization, our findings may be of limited relevance to countries electing to vaccinate with more durable vaccine forms. Finally, the small country size, the organized healthcare system, and the unique ethnic makeup affect the vaccination compliance of the population.

Of note, the vaccine availability in Israel was equal between different municipalities, and vaccination is free to all citizens. When faced with monetary and insurance barriers to healthcare which may exist in other countries, the need to directly target socio-economically disadvantaged populations is even more urgent. Our work, is the first to confirm, in a real-world settings that prior concerns raised regarding vaccination acceptance in lower SES were warranted and should be addressed accordingly. In conclusion, we found a significant correlation between the municipal SES and the vaccination rates of at-risk populations. We also point to a negative correlation between active disease burden and vaccination rates of these populations. As the initial vaccine rollout is limited, case numbers are spiking and more infectious COVID-19 strains are emerging, we urge policymakers to emphasize efforts of vaccination in municipalities with lower SES while using the suggested novel, metric, the VNR, to target the vaccination efforts. Geographic heatmap layering of the VNR can further assist in preventing pockets of regional decreased immunity.

## Data Availability

Data will be available upon request.

Https://www.covid19maps.org

